# Neutralizing Antibodies Responses to SARS-CoV-2 in COVID-19 Inpatients and Convalescent Patients

**DOI:** 10.1101/2020.04.15.20065623

**Authors:** Xiaoli Wang, Xianghua Guo, Qianqian Xin, Yang Pan, Jing Li, Yanhui Chu, Yingmei Feng, Quanyi Wang

## Abstract

**Background:** COVID-19 is a pandemic with no specific antiviral treatments or vaccines. The urgent needs for exploring the neutralizing antibodies from patients with different clinical characteristics are emerging.

**Methods:** A total of 117 blood samples were collected from 70 COVID-19 inpatients and convalescent patients. The presence of neutralizing antibody was determined with a modified cytopathogenic assay based on live SARS-CoV-2. The dynamics of neutralizing antibody levels at different with different clinical characteristics were analyzed.

**Results:** The seropositivity rate reached up to 100.0% within 20 days since onset, and remained 100.0% till day 41-53. The total GMT was 1:163.7 (95% CI, 128.5 to 208.6), and the antibody level was highest during day 31-40 since onset, and then decreased slightly. Individual differences in changes of antibody levels were observed among 8 representative convalescent patients. In multivariate GEE analysis, patients at age of 31-60 and 61-84 had a higher antibody level than those at age of 16-30 (β=1.0518, *P*=0.0152; β=1.3718, *P*=0.0020). Patients with a worse clinical classification had a higher antibody titer (β=0.4639, *P*=0.0227).

**Conclusions:** The neutralizing antibodies were detected even at the early stage of disease, and a significant response showed in convalescent patients. Moreover, changes on antibody levels ware individual specific.

## Introduction

The family Coronaviridae is comprised of large, enveloped, single-stranded, and positive-sense RNA viruses that can infect a wide range of animals and human ^[1]^. Two coronavirus pandemics in human have emerged in the past two decades. Severe acute respiratory syndrome coronavirus (SARS-CoV) was first recognized in 2003, causing a global outbreak ^[2]^. It was followed by another pandemic event in 2012 designated as Middle East respiratory syndrome coronavirus (MERS-CoV) ^[3]^. In December 2019, emergence of severe acute respiratory syndrome coronavirus 2 (SARS-CoV-2) originating in Wuhan, China, has rapidly spread worldwide, and the World Health Organization (WHO) declared coronavirus disease 2019 (COVID-19) a pandemic. As of April 12, 2020, the cases of COVID-19 have been reported in 211 countries and territories worldwide, with a total of 1696588 confirmed cases and 105952 deaths ^[4]^. Moreover, the number of confirmed cases continues to grow at a rapid rate, including United States ^[5]^. To date, the outbreak in China has been effectively controlled by widespread testing, quarantine of cases, contact tracing and social distancing ^[6]^. As of April 12, 2020, there were only 1156 active cases in China ^[7]^. Despite supportive care and conventional anti-virus therapies, neither antiviral treatments nor vaccines that could specifically target against COVID-19 have been achieved ^[8]^.

Neutralizing antibodies play an important role in virus clearance and have been considered as a key immune product for protection or treatment against viral diseases. The results from some researches indicated that using convalescent plasma on Ebola, SARS-CoV and H5N1 avian influenza patients were proved to be effective ^[9]^, moreover, COVID-19 Joint Investigation Report by China-WHO pointed out that serum collected from COVID-19 convalescent patients can fully neutralize the cellular infectivity of the isolated virus ^[10]^. In addition, Shen et al ^[11]^ pointed out that 5 critically ill patients with COVID-19, administration of convalescent plasma containing neutralizing antibody was followed by improvement in their clinical status. These findings raise the hypothesis that using convalescent plasma transfusion could also be beneficial in COVID-19 patients. However, immunity duration and changes on immunity levels of patients in convalescent period remains largely unknown. Given the knowledge gap of this area, we determined that an updated analysis of antibody levels of COVID-19 patients at different time and severity of illness might help develop rapid diagnostic reagents, vaccines, drugs, and other treatments. It’s of great significance for the long-term control and treatment of COVID-19.

The purpose of this current study was to analyze the dynamics of neutralizing antibody levels at different time since onset from different severity COVID-19 inpatients and convalescent patients, and to provide information for the scientific community to understand, detect, and treat COVID-19.

## Materials and Methods

### Study Design and Subjects

COVID-19 case definition and clinical classification based on severity were defined according to the New Coronavirus Pneumonia Prevention and Control Protocol for the novel coronavirus disease 2019 (COVID-19) (7th edition) released by the National Health Commission of China. Seventy COVID-19 patients were enrolled from A hospital and B hospital, of whom 12 were inpatients and 58 were convalescent patients. To study the kinetics of neutralizing antibody production, blood samples were collected from two, three and four time points in 19, 8 and 4 patients, respectively. Together with 39 patients with only one blood sample collection, totaled 117 blood samples were analyzed in the study. The protocol of the study was reviewed and approved by the Medical Ethical Committee of Beijing Youan Hospital, Capital Medical University (approval number LL-2020-041-K). Before enrollment, written informed consent was obtained from each enrolled patient.

To study longitudinal changes of antibody titers, 8 convalescent patients were selected from these 70 patients, including 4 in mild group and 4 in moderate group. Two patients were tested for twice, 2 patients were tested for three time, and 4 patients for four times.

### Clinical Measurements

The patients’ anthropometric characteristics and information on gender, age, medical history, smoking and drinking habits, and intake of medications were collected. In addition, the history of residence in or traveling to Wuhan in recent weeks was obtained. White blood cell and different white blood cell count were obtained.

### Immunogenicity Assessment

The indicators for immunogenicity assessment included seropositivity rate and determination of the geometric mean titer (GMT). The neutralizing antibody titer was calculated by Reed-Muench method on day 5. A titer of 1:4 or higher indicated seropositivity. For calculation of GMT, antibody titers of <1:8, >1:512, and >1:1024 were assigned values of 1:4, 1:(512+512/2), and 1:(1024+1024/2), respectively.

The presence of neutralizing antibody was determined with a modified cytopathogenic assay. Serum samples were inactivated at 56°C for 30 minutes and serially diluted with cell culture medium in two-fold steps. The diluted serums were mixed with a virus suspension of 100 CCID50 in 96-well plates at a ratio of 1:1, followed by 2 hours incubation at 36.5°C in a 5% CO_2_ incubator. 1-2 X104 Vero cells were then added to the serum-virus mixture, and the plates were incubated for 5 days at 36.5°C in a 5% CO_2_ incubator. Cytopathic effect (CPE) of each well was recorded under microscopes, and the neutralizing titer was calculated by the dilution number of 50% protective condition.

### Statistical Analysis

Mean with standard deviation was used for continuous variables description, and number with percentage was used for categorical variables description. Median with minimum and maximum was used to describe days for antibody testing of 1st sample since onset. Kruskal-Wallis rank-sum nonparametric method was used to compare log-transformed neutralizing antibody values. The comparison of categorical data was performed using Chi-square test or Fisher’s exact test. The association between antibody levels and potential factors, i.e., gender, age, clinical classification, and time since onset of symptoms, were estimated by Generalized Estimating Equations (GEE) model with logit link function, which took into account the correlation between repeated measurements of each patient. Hypothesis testing was two-sided with an alpha value of 0.05. Analyses were conducted by SAS 9.4 (SAS Institute, Cary, NC, USA).

## Results

### Characteristics of the Patients

Of the 70 patients enrolled into this study, 58 were recovered and discharged from hospital (Table 1). The mean age of the patients was 45.1 years (range 16.0-84.0). A total of 58.6% were female. Among them, 38 (54.2%) patients were residents or travelled in Wuhan, Hubei. The number of patients having history of cardiovascular disease, diabetes, and hypertension was 2 (2.8%), 5 (7.1%) and 9 (12.9%), respectively. One (1.4%) patient was asymptomatic infected, 22 (31.4%) had mild clinical manifestations, 43 (61.5%) were moderate, and the remaining 4 (5.7%) were in severe condition (1 was inpatients and 3 were convalescent patients). Circulating C-reactive protein for inpatients and convalescent patients were 7.5 and 17.2 mg/L, respectively. For the neutralizing antibody test of 1^st^ sample since onset in this study, the median time was 33.0 days (range 10.0-53.0), and the time of convalescent patients (35.0 days) were longer than inpatients (13.5 days).

**Table 1.** Demographics and clinical characteristics of the COVID-19 patients.

| Characteristic | Total | Inpatients | Convalescent patients |
| --- | --- | --- | --- |
| Number of patients | 70 | 12 | 58 |
| Male, n (%) | 29 (41.4%) | 4 (33.3%) | 27 (43.1%) |
| History of residence or traveling in Wuhan, n(%) | 38 (54.2%) | 5 (41.7%) | 33 (56.9%) |
| History of cardiovascular disease, n(%) | 2 (2.8%) | 0 | 2 (3.4%) |
| History of diabetes, n(%) | 5 (7.1%) | 0 | 5 (8.6%) |
| History of hypertension, n(%) | 9 (12.9%) | 0 | 9 (15.5%) |
| Clinical classification, n(%) |  |  |  |
| Asymptomatic | 1 (1.4%) | 0 (0.0%) | 1 (1.7%) |
| Mild | 22 (31.4%) | 3 (25.0%) | 19 (32.8%) |
| Moderate | 43 (61.5%) | 8 (66.7%) | 35 (60.3%) |
| Severe | 4 (5.7%) | 1 (8.3%) | 3 (5.2%) |
| Age(years) | 45.1±14.2 | 42.7±11.6 | 45.6±14.7 |
| Body temperature at admission (°C) | 37.1±0.7 | 37.0±0.7 | 37.1±0.7 |
| Systolic blood pressure (mm Hg) | 126.1±16.9 | 130.2±8.4 | 125.2±17.9 |
| Diastolic blood pressure (mm Hg) | 78.4±12.4 | 81.5±10.0 | 77.7±2.8 |
| White blood cells (10<sup>9</sup>/L) | 4.3±1.4 | 4.1±1.4 | 4.4±1.4 |
| Neutrophil (10<sup>9</sup>/L) | 2.6±1.2 | 2.5±1.2 | 2.6±1.2 |
| Lymphocytes (10<sup>9</sup>/L) | 1.3±0.6 | 1.3±0.5 | 1.3±0.6 |
| Monocytes (10<sup>9</sup>/L) | 0.3±0.1 | 0.3±0.1 | 0.3±0.1 |
| Platelets (10<sup>12</sup>/L) | 202.0±70.8 | 169.2±50.2 | 209.4±72.4 |
| Circulating C-reactive protein (mg/L) | 15.3±20.1 | 7.5±11.8 | 17.2±22.1 |
| Days for antibody testing of 1<sup>st</sup> sample since onset, median (min, max) | 33.0 (10.0, 53.0) | 13.5 (10.0, 22.0) | 35.0 (12.0, 53.0) |

Data are expressed as mean±SD if not specified.

### Changes on Antibody Levels with Days since Onset

The seropositivity rate reached up to 100.0% for 117 blood samples at different stages of illness. The total GMT was 1:163.7 (95% CI, 128.5 to 208.6), of which 52.1% (61/117) had a titer between 1:64 and 1:512 (Table 2).

**Table 2.**
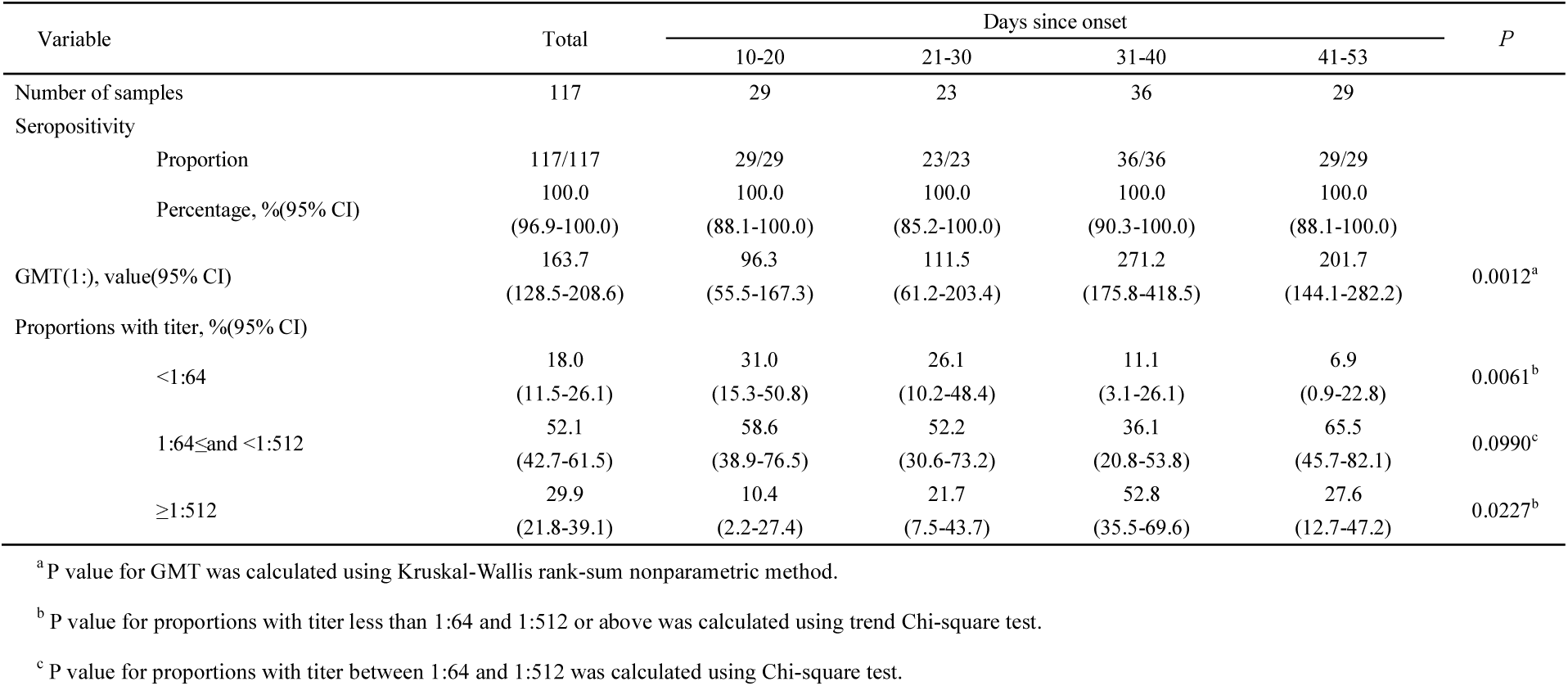
Seropositivity rates and antibody levels in 117 blood samples at different time since onset.

| Variable | Total | Days since onset |  |  |  | *P* |
| --- | --- | --- | --- | --- | --- | --- |
|  |  | 10-20 | 21-30 | 31-40 | 41-53 |  |
| Number of samples | 117 | 29 | 23 | 36 | 29 |  |
| Seropositivity |  |  |  |  |  |  |
| Proportion | 117/117 | 29/29 | 23/23 | 36/36 | 29/29 |  |
| Percentage, %(95% CI) | 100.0<br>(96.9-100.0) | 100.0<br>(88.1-100.0) | 100.0<br>(85.2-100.0) | 100.0<br>(90.3-100.0) | 100.0<br>(88.1-100.0) |  |
| GMT(1:), value(95% CI) | 163.7<br>(128.5-208.6) | 96.3<br>(55.5-167.3) | 111.5<br>(61.2-203.4) | 271.2<br>(175.8-418.5) | 201.7<br>(144.1-282.2) | 0.0012<sup>a</sup> |
| Proportions with titer, %(95% CI) |  |  |  |  |  |  |
| <1:64 | 18.0<br>(11.5-26.1) | 31.0<br>(15.3-50.8) | 26.1<br>(10.2-48.4) | 11.1<br>(3.1-26.1) | 6.9<br>(0.9-22.8) | 0.0061<sup>b</sup> |
| 1:64≤and <1:512 | 52.1<br>(42.7-61.5) | 58.6<br>(38.9-76.5) | 52.2<br>(30.6-73.2) | 36.1<br>(20.8-53.8) | 65.5<br>(45.7-82.1) | 0.0990<sup>c</sup> |
| ≥1:512 | 29.9<br>(21.8-39.1) | 10.4<br>(2.2-27.4) | 21.7<br>(7.5-43.7) | 52.8<br>(35.5-69.6) | 27.6<br>(12.7-47.2) | 0.0227<sup>b</sup> |

<sup>a</sup> P value for GMT was calculated using Kruskal-Wallis rank-sum nonparametric method.
<sup>b</sup> P value for proportions with titer less than 1:64 and 1:512 or above was calculated using trend Chi-square test.
<sup>c</sup> P value for proportions with titer between 1:64 and 1:512 was calculated using Chi-square test.

We analyzed the changes on antibody levels according to the time course since onset (Table 2). The antibody levels according to different time course since onset were significantly different (*P*=0.0012). The GMT of day 31-40 since onset (1: 271.2, 95% CI, 175.8 to 418.5) reached the highest, and decreased slightly after that time period. Univariate GEE analysis showed that the antibody level during day 31-40 was significantly higher than other phases (Table 4). However, multivariate GEE analysis showed that the antibody level during day 31-40 was only higher than day 10-20 (β= -0.6276, *P*=0.0201).

**Table 3.** Antibody levels in 70 patients by gender, age, clinical classification, and recovery.

| Characteristic |  | GMT* (1:), value(95% CI) | P<sup>#</sup> |
| --- | --- | --- | --- |
| Gender | Male | 168.6(101.2-280.9) | 0.7243 |
|  | Female | 185.6(129.1-266.6) |  |
| Age(years) | 16-30 | 71.0(27.7-181.8) | 0.0140 |
|  | 31-60 | 200.6(150.4-267.6) |  |
|  | 61-84 | 220.1(71.8-674.8) |  |
| Clinical classification | Asymptomatic or mild | 141.9(79.5-253.2) | 0.2435 |
|  | Moderate or severe | 199.5(141.8-280.5) |  |
| Recovery or not | Inpatients | 76.1(33.5-172.9) | 0.0055 |
|  | Convalescent patients | 212.7(157.5-287.3) |  |

\*The geometric mean of repeated measurements for each patient was used to represent the only testing result.
<sup>#</sup>P value for GMT was calculated using Kruskal-Wallis rank-sum nonparametric method.

**Table 4.** Univariate and multivariate GEE analysis of factors associated with antibody levels.

| Characteristic |  | Univariate | Multivariate |  |  |  |
| --- | --- | --- | --- | --- | --- | --- |
|  |  | *P* | Coefficient | Standard error | 95% CI | *P* |
| Gender | Female vs. male | 0.9426 | 0.0423 | 0.2304 | -0.4092-0.4938 | 0.8543 |
| Age(years) |  |  |  |  |  |  |
|  | 31-60 vs. 16-30 | 0.0164 | 1.0518 | 0.4334 | 0.2025-1.9012 | 0.0152 |
|  | 61-84 vs. 16-30 | 0.0061 | 1.3718 | 0.4447 | 0.5002-2.2434 | 0.0020 |
| Clinical classification | Moderate or severe vs. mild | 0.0753 | 0.4639 | 0.2036 | 0.0649-0.8630 | 0.0227 |
| Days since onset |  |  |  |  |  |  |
|  | 10-20 vs. 31-40 | 0.0284 | -0.6276 | 0.2700 | -1.1569- -0.0983 | 0.0201 |
|  | 21-30 vs. 31-40 | 0.0410 | -0.5152 | 0.2765 | -1.0571-0.0267 | 0.0624 |
|  | 41-53 vs. 31-40 | 0.0152 | -0.2075 | 0.2616 | -0.7203-0.3053 | 0.4277 |

Blood samples at different time since onset also showed differences in the distribution of neutralizing antibody titers (Table 2 and Figure 1). The proportion with a titer less than 1:64 decreased with days since onset (*P*_trend_=0.0061), and the lowest was found during day 41-53. During day 41-53 since onset, there were 65.5% of samples with a titer between 1:64 and 1:512, not significantly different from other phases (*P*=0.0990). The proportion with a titer of 1:512 or above increased with days since onset (*P*_trend_=0.0227), and peaked the highest during day 31-40.

**Figure 1.**
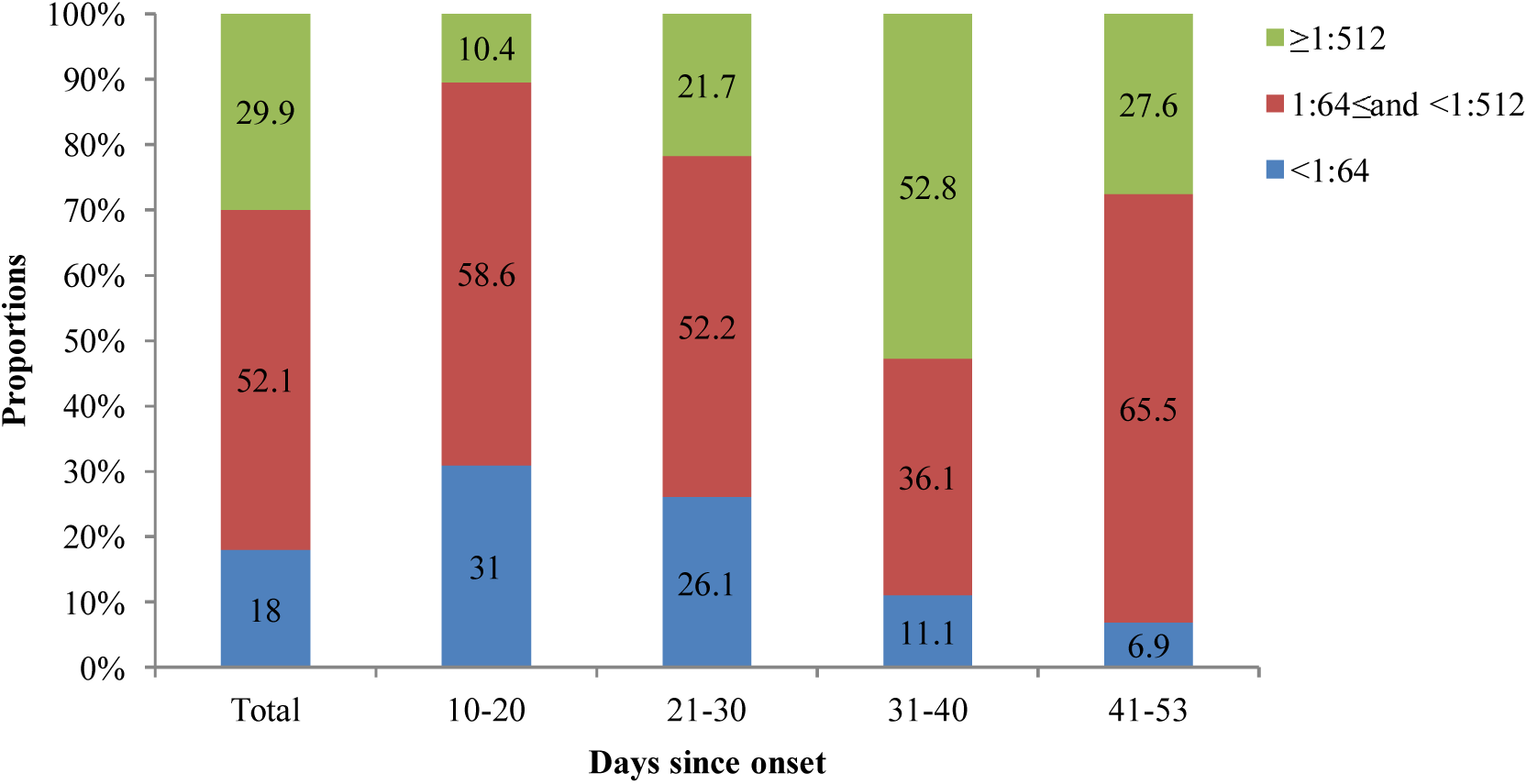
Distribution of neutralizing antibody titers in 70 patients at different time since onset.

### Dynamics of Antibody Titers in Representative Convalescent Patients since Onset

Among the 8 convalescent patients, days of neutralizing antibody tests since onset ranged from 12.0 to 60.0. During day 12-25, antibody titers of 4 patients (c, d, e, and f) were on an increasing curve, however, 4 patients (a, b, g, and h) were on a declining curve (Figure 2). Then during day 26-60, antibody titers showed a marked increase in 4 patients (a, b, c, and d) but a decrease in 3 patients (e, f, and h). One patient (g) remained a stable titer of 1:128. It should be noted that the antibody titer of 1 patient (f) decreased from 1:1536 on day 20 to 1:48 on day 43.

**Figure 2.**
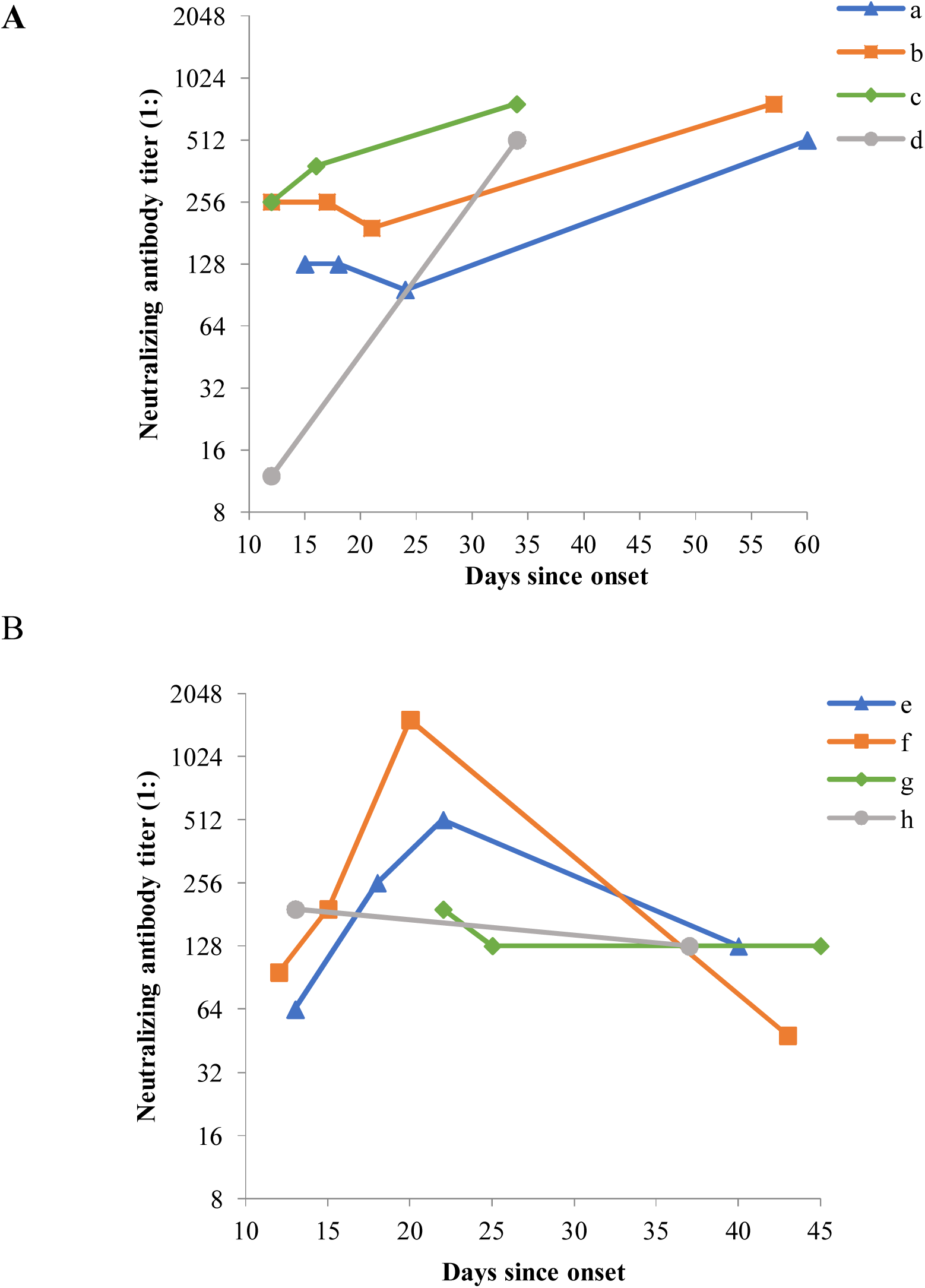
Dynamics of neutralizing antibody titers in 8 convalescent COVID-19 patients since onset. In the figure, a, b, c, d, e, f, g and h represented the 8 representative convalescent patients. In Figure 2-A, four patients whose antibody titers showed the obvious increasing trend were included. Figure 2-B included the other four patients whose antibody titers showed the decreasing trend.

### Changes on Antibody Levels with Clinical Classification

The neutralizing antibody titers were similar in the two gender groups, of which 1:168.6 (95% CI, 101.2 to 280.9) in male and 1:185.6 (95% CI, 129.1 to 266.6) in female. The effect of gender was also not statistically significant in both univariate (*P*=0.9426) and multivariate (*P*=0.8543) GEE analysis. A significant neutralizing antibody response was observed in older patients with a geometric mean titer of 1:220.1 (95% CI, 71.8 to 674.8) compared to patients at age of 16-30 (1:71.0, 95% CI, 27.7 to 181.8) and at age of 31-60 (1:200.6, 95% CI, 150.4 to 267.6) (*P*=0.0140). In multivariate GEE analysis, patients at age of 31-60 and 61-84 were more likely to have a higher antibody level than those at age of 16-30 (β=1.0518, *P*=0.0152; β=1.3718, *P*=0.0020).

Compared to the patients with asymptomatic or mild manifestations (GMT 1:141.9, 95% CI, 79.5 to 253.2), the antibody levels were similar to patients with moderate or severe condition (GMT 1:199.5, 95% CI, 141.8 to 280.5). However, after adjusting other factors, patients with more severe symptoms tended to have a higher antibody titer (β=0.4639, *P*=0.0227). The GMT of convalescent patients was 1:212.7(95% CI, 157.5 to 287.3), and was higher than inpatients (1:76.1, 95% CI: 33.5 to 172.9; *P*=0.0055). Details were listed in Table 3 and 4.

## Discussion

Due to the COVID-19 widely spreading around the world, the specific therapeutic agents or vaccines for COVID-19 are urgently needed. We analyzed the neutralizing antibody levels among different classification patient with increased days since onset of symptoms, the results indicated that a significant neutralizing antibody response was observed in convalescent patients, moreover, there were individual differences in changes of antibody levels.

In the study, typical antibody responses to live viral infection were seen induced in all COVID-19 patients regardless the stage of the disease. Moreover, the immunity levels were already strong enough even at the early stage of illness, with seropositivity rate reaching up to 100.0% at day 10. The GMT reached the peak at day 31-40 after onset of symptoms. Even though the GMT had a slightly decreased at day 41-53, the seropositivity rate remained 100.0%. However, the results were different from other study which indicated the titers of antibodies reached their peak at 10 to 15 days after disease onset ^[12]^. After adjusting cofounding factors, multivariate GEE analysis demonstrated that the antibody levels were comparable between day 31-40 and day 41-53 since disease onset. However, the proportion with a titer of 1:512 or above decreased from 52.8% during day 31-40 to 27.6% during day 41-53. Due to the lack of blood samples collected from patients in the later stages of illness, whether and when seropositivity rate declined below 100.0% remained unknown. How long any immunity in patients will last is a key and big unknown problem for safe and effect antiviral treatments and vaccines in the future ^[13]^. For other coronaviruses, immunity after an infection was strong for several months, but then began to wane ^[14]^. Chang ^[15]^ indicated that peak IgG titers in all SARS-CoV patients appeared at 1 month or 1 to 3 months after the disease onset, and a drop (4.4-fold on average) in IgG titers was evident between 1 month and 6 months after onset. These studies suggested that it’s worthy of further study to analyze antibodies after COVID-19 patients recovered for a longer time.

Notably, when antibody titers of 4 representative patients continued to increase after 25 days since onset, 1 patient kept unchanged at a low antibody titer. However, other 3 patients started to decrease. In addition, most of patients had middle titer levels, while 6.9% (2/29) patients with a low titer of less than 1:64 was still screened out during day 41-53 since onset, suggesting whether these patients were at high risk of reinfection was unclear and should be explored in further. These results fully supported the speculation about individual specificity, which should be paid attention to in the diagnosis and treatment of COVID-19 patients. It’s necessary to explore individual specificity of changes on antibody levels with more subjects and specially designed trials.

When comparing antibody levels between gender, age, and clinical classification, it should be noted that the GMTs were not practical testing values, so they were only used to imply which group were on the high level. Male and female were on the similar antibody level, nevertheless, we found that the neutralizing antibody titers of the 70 patients significantly increased with age. Wu ^[12]^ also showed that elderly and middle-age COVID-19 patients had significantly higher plasma antibody titers and spike-binding antibodies than young patients. High antibody levels might result from strong immune response against SARS-CoV-2 in these elderly patients. Whether high antibody levels protect these patients from progression into severe or critical conditions deserved further studies.

The results of our study indicated that convalescent patients had a higher antibody level than inpatients, which highlight the positive correlation between recovery and days since onset (Spearman correlation coefficient=0.5426, *P*<0.0001). Besides, we also found that antibody levels in asymptomatic or mild patients were slightly lower than moderate or severe patients, which was in line with other previous studies ^[16-17]^. Zhang ^[17]^ came to a similar conclusion that severe cases were more frequently found in COVID-19 patients with high IgG levels, compared to those who with low IgG levels. Previous data showed that severe SARS-CoV was also associated more robust serological responses including early seroconversion and higher IgG levels ^[18-19]^. Therefore, antibodies against SARS-CoV-2 increased with the upgrading of clinical classification.

Several limitations of this study should be noted. First, the involved patients were selected by convenient sampling instead of random sampling. So the representativeness is relatively insufficient, and the samples could only represent the general situation to a certain extent. Second, among 70 patients, only 12 of them were followed up more than twice, and the average follow-up period was relatively short, about only 14.3 days (range 3.0-36.0). Therefore, it is of value to follow up patients for a longer time. Third, the subjects were mainly mild or moderate by illness severity, and only 1 asymptomatic patient and 4 severe patients, whether a different antibody levels would have been observed with different severity patients cannot be determined. Hence, it’s important to evaluate antibody levels in asymptomatic infected and critical patients in future study.

In conclusion, this study showed that all COVID-19 patients were seropositivity to SARS-CoV-2 even at the early stage of illness, and a significant neutralizing antibody response was observed in convalescent patients. Neutralizing antibody levels increased with days since onset of symptoms, elder age and the worsening of clinical classification. Changes on antibody levels ware individual specific.

## Data Availability

All data referred to in the manuscript were from our own testing. 

## Notes

### Contributions

Concept and design: Yingmei Feng and Quanyi Wang.

Acquisition, analysis, or interpretation of data: All Authors.

Drafting of the manuscript: Xiaoli Wang, Xianghua Guo, and Qianqian Xin.

Critical revision of the manuscript: Yingmei Feng and Quanyi Wang.

Laboratory testing: Jing Li

Data management and statistical analysis: Yanhui Chu and Yang Pan

## Acknowledgments

We thank all local health workers and local CDC professionals for their contributions to field investigation.

## References

[1] Huang JM, Jan SS, Wei XB, et al. Evidence of the Recombinant Origin and Ongoing Mutations in Severe Acute Respiratory Syndrome 2 (SARS-COV-2). bioRxiv 2020; published online Mar 17. DOI: 10.1101/2020.03.16.993816.

[2] Zhong N. Management and Prevention of SARS in China. hilosophical Transactions of the Royal Society B Biological Sciences. 2004; 359(1447):1115–1116. DOI: 10.1098/rstb.2004.1491.

[3] Lu L, Liu, Q, Du L, et al. Middle East Respiratory Syndrome Coronavirus (MERS-CoV): Challenges in Identifying Its Source and Controlling Its Spread. Microbes Infect. 2013; 15:625–629. DOI: 10.1016/j.micinf.2013.06.003.

[4] WHO. Coronavirus Disease (COVID-2019) Situation Reports. Apr 12, 2020. https://www.who.int/emergencies/diseases/novel-coronavirus-2019/situation-reports (accessed Apr 12, 2020).

[5] Liu QH, Zhu JK, Liu ZC, et al. Assessing the Global Tendency of COVID-19 Outbreak. medRxiv 2020; published online Mar 18. DOI: 10.1101/2020.03.18.20038224.

[6] The Lancet. COVID-19: Learning from Experience. Lancet. 2020; 395:1011. DOI: 10.1016/S0140-6736(20)30686-3.

[7] National Health Commission of China. Do Our Best to Prevent and Control the Outbreak of the New Type of Tubular Virus Pneumonia: Report on Situation. Apr 12, 2020. http://www.nhc.gov.cn/xcs/yqtb/202004/fa7bb40a7fbf4b2c8f3989d512fe5b77.shtml (accessed Apr 12, 2020). (In Chinese)

[8] Shang WL, Yang Y, Rao YF, et al. The Outbreak of SARS-CoV-2 Pneumonia Calls for Viral Vaccines. npj Vaccines. 2020; 5:18. DOI: 10.1038/s41541-020-0170-0.

[9] Chen L, Xiong J, Ba L, et al. Convalescent Plasma as a Potential Therapy for COVID-19. Lancet. 2020; 20:398–400. DOI: 10.1016/S1473-3099(20)30141-9.

[10] Bureau of Disease Control and Prevention, National Health Commission of China. COVID-19 Joint Investigation Report by China and WHO. http://www.nhc.gov.cn/jkj/s3578/202002/87fd92510d094e4b9bad597608f5cc2c.shtml (accessed Apr 9, 2020). (In Chinese)

[11] Li DL, Yang MH, Xing L, et al. Treatment of 5 Critically Ill Patients With COVID-19 With Convalescent Plasma. JAMA 2020; published online Mar 27. DOI: 10.1001/jama.2020.4783.

[12] Wu F, Wang AJ, Liu M, et al. Neutralizing Antibody Responses to SARS-CoV-2 in a COVID-19 Recovered Patient Cohort and their Implications. medRxiv 2020; published online Mar 30. DOI: 10.1101/2020.03.30.20047365.

[13] Ewen Callaway. Coronavirus Vaccines: Five Key Questions as Trials Begin. Nature News Explainer. 2020; 579:481. https://media.nature.com/original/magazine-assets/d41586-020-00798-8/d41586-020-00798-8.pdf.

[14] Fatima Amanat, Thi Nguyen, Veronika Chromikova, et al. A Serological Assay to Detect SARS-CoV-2 Seroconversion in Humans. medRxiv 2020; published online Mar 18. DOI: 10.1101/2020.03.17.20037713.

[15] Chang SC, Wang JT, Huang LM, et al. Longitudinal Analysis of Severe Acute Respiratory Syndrome (SARS) Coronavirus-specific Antibody in SARS Patients. Clin Diagn Lab Immunol. 2005; 12(12):1455–1457. DOI: 10.1128/CDLI.12.12.1455-1457.2005.

[16] Zhao JJ, Yuan Q, Wang HY, et al. Antibody Responses to SARS-CoV-2 in Patients of Novel Coronavirus Disease 2019. medRxiv 2020; published online Mar 2. DOI: 10.1101/2020.03.02.20030189.

[17] Zhang BC, Zhou XY, Zhu CL, et al. Immune Phenotyping Based on Neutrophil-to-lymphocyte Ratio and IgG Predicts Disease Severity and Outcome for Patients with COVID-19. medRxiv 2020; published online Mar 12. DOI: 10.1101/2020.03.12.20035048.

[18] Lee N, Chan PK, Ip M, et al. Anti-SARS-CoV IgG Response in Relation to Disease Severity of Severe Acute Respiratory Syndrome. J Clin Virol. 2006; 35:179–184. DOI: 10.1016/j.jcv.2005.07.005.

[19] Zhang L, Zhang F, Yu W, et al. Antibody Responses against SARS Coronavirus are Correlated with Disease Outcome of Infected Individuals. J Med Virol. 2006; 78:1–8. DOI: 10.1002/jmv.20499.

